# Parakinesia Brachialis Oscitans and Post-Stroke Motor Recovery: A Propensity Score-Matched Cohort Study

**DOI:** 10.64898/2026.01.29.26345175

**Authors:** Congcong Wang, Runying Wang, Hua Hu, Xiaojun Tian, Shuangxi Guo, Zhou Su

## Abstract

**Objective:** A comparative analysis was conducted on the rehabilitation effects of limb functions in patients with post-stroke yawning-induced parakinesia brachialis oscitansysis (PBO), patients without PBO, and patients whose PBO naturally disappeared after the onset of the disease.

**Methods:** The study included ischemic stroke patients diagnosed and treated in our hospital from March 2024 to June 2024. Patients were divided into two groups: the PBO group and the non-PBO group, based on whether PBO was administered. Propensity score matching was employed to account for all covariates and perform a 1:2 matching to balance the baseline characteristics of the two groups. The matched data were used for subsequent analysis to observe the Lovett scores and FMA scores of the two groups 3 months after the onset. For 33 patients with PBO, they were divided into two groups: the persistent group and the disappearing group, based on whether the PBO lasted for more than 1 month. The Lovett scores and FMA scores of the two groups were observed 3 months after the onset.

**Results:** After propensity score matching, there were 26 patients in the PBO group and 52 patients in the non-PBO group. The baseline characteristics of the two groups were basically balanced, and the difference was not statistically significant (P>0.05). Compared with the non-PBO group, the Lovett scores and FMA scores of the PBO group 3 months after the onset were higher, and the difference was statistically significant (P < 0.05). Compared with the PBO persistent group, the FMA score of the PBO disappearing group 3 months after the onset was higher than that of the persistent group, and the difference was statistically significant (P < 0.05). There was no statistically significant difference in Lovett muscle strength between the two groups (P > 0.05).

**Conclusion:** The functional recovery of patients with PBO was better than that of patients without PBO manifestation 3 months after the initial diagnosis. Moreover, patients whose PBO appeared first and then disappeared had better functional recovery than those whose PBO persisted.

## Introduction

Ischemic stroke is the most common disabling disease in China. With the acceleration of population aging, its incidence continues to rise, with over 3 million new cases annually^[1]^. The resulting loss of working capacity imposes significant life and economic burdens on patients and their families. Early rehabilitation therapy, particularly limb function rehabilitation, significantly impacts patients’ subsequent quality of life^[2]^. Parakinesia brachialis oscitans (PBO) refers to the phenomenon where paralyzed limbs exhibit involuntary movement during yawning, observed after neurological damage causes loss of voluntary limb movement. This uncommon occurrence was first termed PBO by Walusinski in 2005^[3]^. Current research on PBO, both domestically and internationally, remains limited to scattered case reports and mechanistic hypotheses. Systematic, multimodal explanations of its mechanisms are lacking, and high-level, evidence-based medical evidence linking PBO to rehabilitation prognosis is absent^[4]^. Clinically, rehabilitation outcomes for patients exhibiting PBO appear distinct from those without this phenomenon. Against this backdrop, we analyzed short- and long-term functional recovery in ischemic stroke patients with PBO, aiming to enhance limb function rehabilitation by either antagonizing or promoting this phenomenon.

## Methods

### 1. Population

Patients diagnosed with and treated for ischemic stroke at our hospital between March and June 2024 were selected. Inclusion criteria: ① Age 55–75 years; ② The diagnostic workup and therapeutic management of all patients were in accordance with the recommendations outlined in the 2021 AHA/ASA Guideline for the Prevention of Stroke in Patients With Stroke and Transient Ischemic Attack^[5]^; ③ Confirmed acute cerebral infarction via cranial MRI-DWI. ④ Informed consent obtained from patients. Exclusion Criteria: ① Severe psychiatric disorders or cognitive impairment preventing follow-up compliance; ② Unstable cardiovascular disease or poorly controlled chronic conditions; ③ Concurrent fractures or skeletal muscle injuries impairing motor function. Dropout Criteria: ① Patient or family refusal to participate; ②Development of new conditions affecting motor function during the study period, such as fractures. A total of 636 patients meeting these criteria were screened. Patients with documented PBO manifestations in the database were assigned to the experimental group (n=33). Baseline characteristics were described for the 33 PBO patients and 636 patients without PBO manifestations. Propensity Score Matching (PSM) was used to select matched patients from the non-PBO stroke cohort, achieving a 1:2 matching ratio. This study was approved by the Ethics Committee of the First Affiliated Hospital of Xinxiang Medical University (EC-024-588).

### 2 Data Collection

#### 2.1 Baseline Data Collection

Recorded for all subjects: gender, age, stroke location (anterior/posterior circulation), hypertension, diabetes, homocysteine (HCY), and low-density lipoprotein (LDL). Early ischemic changes in the anterior circulation were quantitatively assessed using the Alberta Stroke Program Early CT Score (ASPECTS)^[6]^. For posterior circulation (vertebrobasilar artery system) infarction, evaluation was performed using the posterior circulation ASPECTS (pC-ASPECTS)^[7]^.

#### 2.2 Record the first occurrence time and disappearance time of PBO, or whether the PBO phenomenon persists. Classify cases based on whether the PBO duration exceeds one month into the disappearance group and the persistent group

#### 2.3 Lovett scores

Immediately upon admission and at 3 months, the Lovett scores scale was used to assess muscle strength in the upper and lower limbs affected by the current stroke event. Muscle strength was categorized into grades 0 to 5: Grade 0: No observable muscle contraction; Grade 1: Slight muscle contraction, but unable to produce joint movement; Grade 2: Poor muscle strength, i.e., able to perform a full range of motion at the horizontal plane; Grade 3: Fair strength, allowing full range of motion against gravity but not against resistance; Grade 4: Good strength, allowing movement against moderate resistance; Grade 5: Normal strength, allowing movement against gravity and sufficient resistance.

#### 2.4 The Fugl-Meyer Assessment

(FMA) was administered before and after intervention to evaluate motor function in the upper and lower limbs affected by the current stroke event. This scale comprises two sections: upper limb and lower limb. The upper limb section assesses four domains—arm, wrist, hand, and coordination— with each item scored 0–2 points, yielding a maximum total of 66 points. The lower limb section comprises three domains: supine position, sitting position, and standing position. Each item is scored from 0 to 2 points, with a maximum score of 34 points. Higher scores indicate better motor function^[8].^

#### 2.5 Propensity Score Matching (PSM)

This study employed R 4.3.2 (2023-10-31 ucrt, x86_64-w64-mingw32/x64 64-bit) to construct the matching model. The MatchIt package (version 4.5.5) was used for 1:2 nearest-neighbor propensity score matching without replacement, with a clamp value set at 0.02. The dependent variables were defined as the occurrence (PBO) versus non-occurrence (no PBO) of PBO. Independent variables included gender, age, hypertension, diabetes, hyperlipidemia, homocysteine levels, stroke location, ASPECT score, Lovett score, and FMA score. Unmatched data were excluded, and subsequent analyses were performed on the matched sample.

### Statistical Analysis

Data analysis was performed using SPSS 23.0 statistical software. Quantitative data were first subjected to Shapiro-Wilk normality and Levene’s homogeneity of variance tests. If both criteria were met, data were expressed as x ± s, and intergroup comparisons were conducted using the independent samples t-test. If either criterion was not met, data were expressed as M (P25, P75), and intergroup comparisons were performed using the Mann-Whitney U test. Ordinal data were expressed as M (P25, P75), and intergroup comparisons were performed using the Mann-Whitney U test. Count data were expressed as n (%), and intergroup comparisons were performed using the chi-square test. All tests were two-sided, and P < 0.05 was considered statistically significant.

## Result

1. Comparison of Baseline Data Between the Two Patient Groups Before and After Matching.

The differences in Lovett grading and FMA scores between the two matched groups were statistically significant (P < 0.05). After matching, no statistically significant differences were observed in baseline characteristics between the two groups (P > 0.05), as shown in Table 1.

**Table 1.**
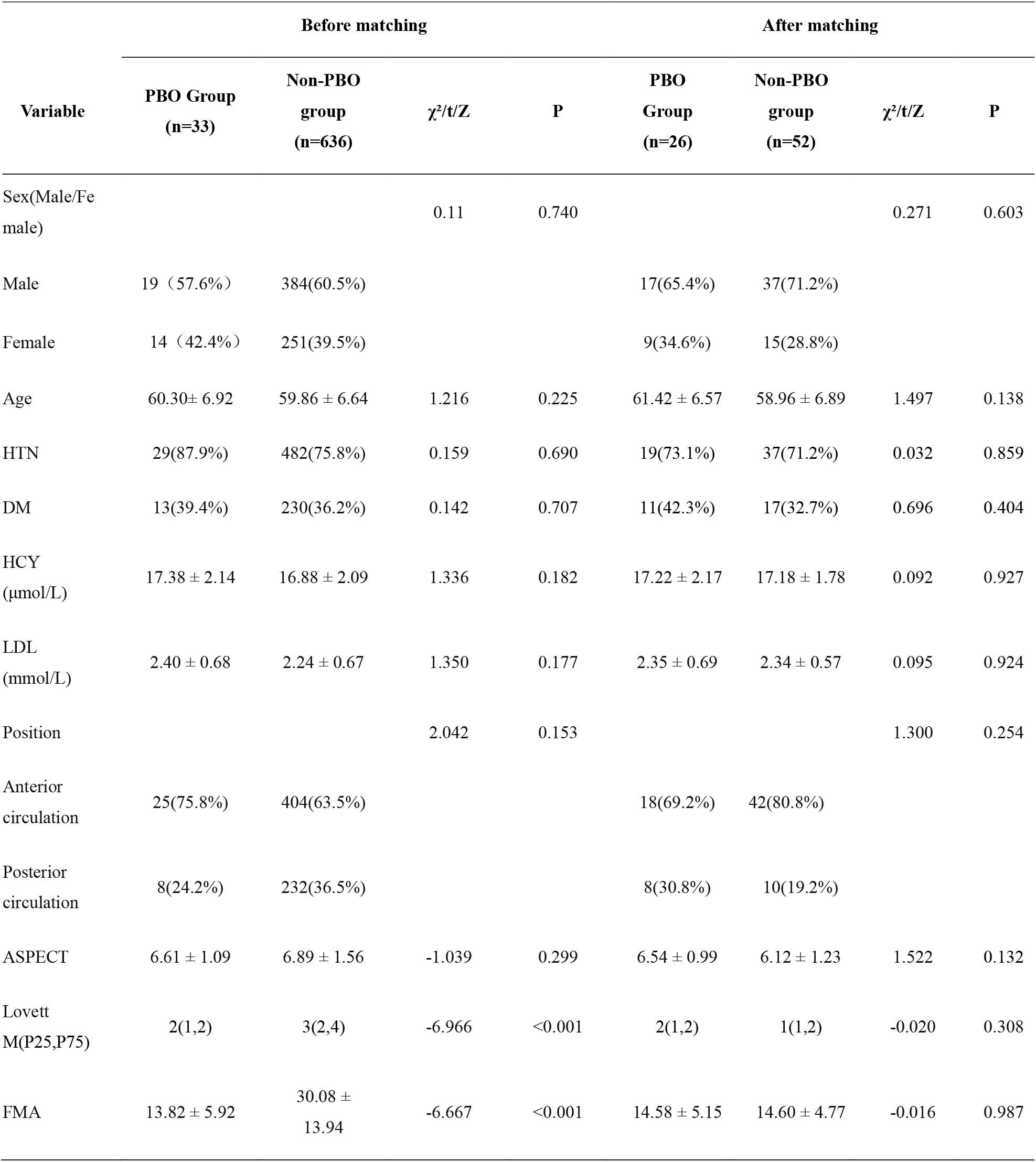
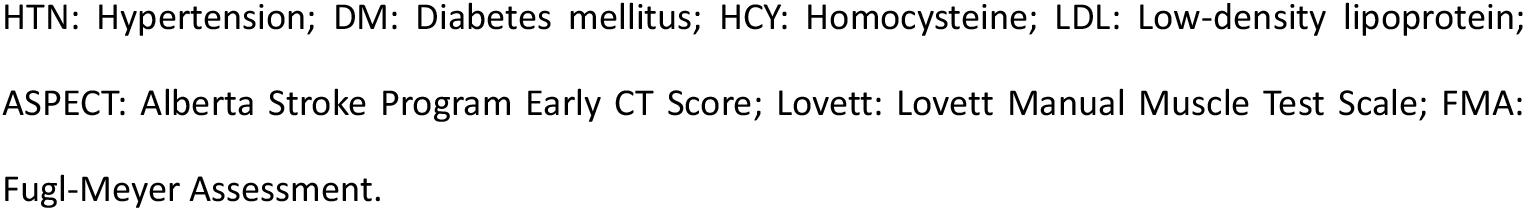
Comparison of Baseline Characteristics Between the Two Patient Groups Before and After Matching.

| Variable | Before matching |  |  |  | After matching |  |  |  |
| --- | --- | --- | --- | --- | --- | --- | --- | --- |
| | PBO Group<br>(n=33) | Non-PBO<br>group<br>(n=636) | $\chi^2/t/Z$ | P | PBO<br>Group<br>(n=26) | Non-PBO<br>group<br>(n=52) | $\chi^2/t/Z$ | P |
| Sex(Male/Fe<br>male) |  |  | 0.11 | 0.740 |  |  | 0.271 | 0.603 |
| Male | 19 (57.6%) | 384(60.5%) |  |  | 17(65.4%) | 37(71.2%) |  |  |
| Female | 14 (42.4%) | 251(39.5%) |  |  | 9(34.6%) | 15(28.8%) |  |  |
| Age | 60.30± 6.92 | 59.86 ± 6.64 | 1.216 | 0.225 | 61.42 ± 6.57 | 58.96 ± 6.89 | 1.497 | 0.138 |
| HTN | 29(87.9%) | 482(75.8%) | 0.159 | 0.690 | 19(73.1%) | 37(71.2%) | 0.032 | 0.859 |
| DM | 13(39.4%) | 230(36.2%) | 0.142 | 0.707 | 11(42.3%) | 17(32.7%) | 0.696 | 0.404 |
| HCY<br>(μmol/L) | 17.38 ± 2.14 | 16.88 ± 2.09 | 1.336 | 0.182 | 17.22 ± 2.17 | 17.18 ± 1.78 | 0.092 | 0.927 |
| LDL<br>(mmol/L) | 2.40 ± 0.68 | 2.24 ± 0.67 | 1.350 | 0.177 | 2.35 ± 0.69 | 2.34 ± 0.57 | 0.095 | 0.924 |
| Position |  |  | 2.042 | 0.153 |  |  | 1.300 | 0.254 |
| Anterior<br>circulation | 25(75.8%) | 404(63.5%) |  |  | 18(69.2%) | 42(80.8%) |  |  |
| Posterior<br>circulation | 8(24.2%) | 232(36.5%) |  |  | 8(30.8%) | 10(19.2%) |  |  |
| ASPECT | 6.61 ± 1.09 | 6.89 ± 1.56 | -1.039 | 0.299 | 6.54 ± 0.99 | 6.12 ± 1.23 | 1.522 | 0.132 |
| Lovett<br>M(P25,P75) | 2(1,2) | 3(2,4) | -6.966 | <0.001 | 2(1,2) | 1(1,2) | -0.020 | 0.308 |
| FMA | 13.82 ± 5.92 | 30.08 ±<br>13.94 | -6.667 | <0.001 | 14.58 ± 5.15 | 14.60 ± 4.77 | -0.016 | 0.987 |

HTN: Hypertension; DM: Diabetes mellitus; HCY: Homocysteine; LDL: Low-density lipoprotein;
ASPECT: Alberta Stroke Program Early CT Score; Lovett: Lovett Manual Muscle Test Scale; FMA:
Fugl-Meyer Assessment.

2. Comparison of Lovett Grading and FMA Between PBO and Non-PBO Groups After 3 Months After 3 months, the PBO group demonstrated significantly higher Lovett grades and FMA scores compared to the non-PBO group (P < 0.05), as shown in Table 2.

**Table 2.**
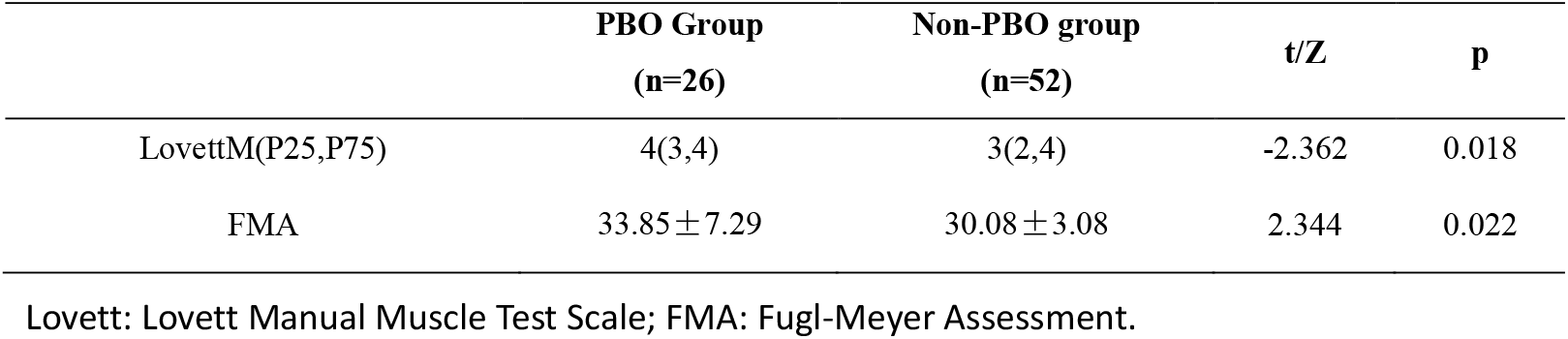
Comparison of Lovett Grading and FMA Between PBO and Non-PBO Groups at 3 Months.

|  | **PBO Group**<br>**(n=26)** | **Non-PBO group**<br>**(n=52)** | **t/Z** | **p** |
| --- | --- | --- | --- | --- |
| LovettM(P25,P75) | 4(3,4) | 3(2,4) | -2.362 | 0.018 |
| FMA | 33.85 ± 7.29 | 30.08 ± 3.08 | 2.344 | 0.022 |

Lovett: Lovett Manual Muscle Test Scale; FMA: Fugl-Meyer Assessment.

2. Comparison of Baseline Data Between the PBO Disappearance Group and the PBO Persistence Group

Comparison of baseline data between the PBO discontinuation group and the PBO continuation group showed no statistically significant differences between the two groups (P > 0.05), as shown in Table 3.

**Table 3.**
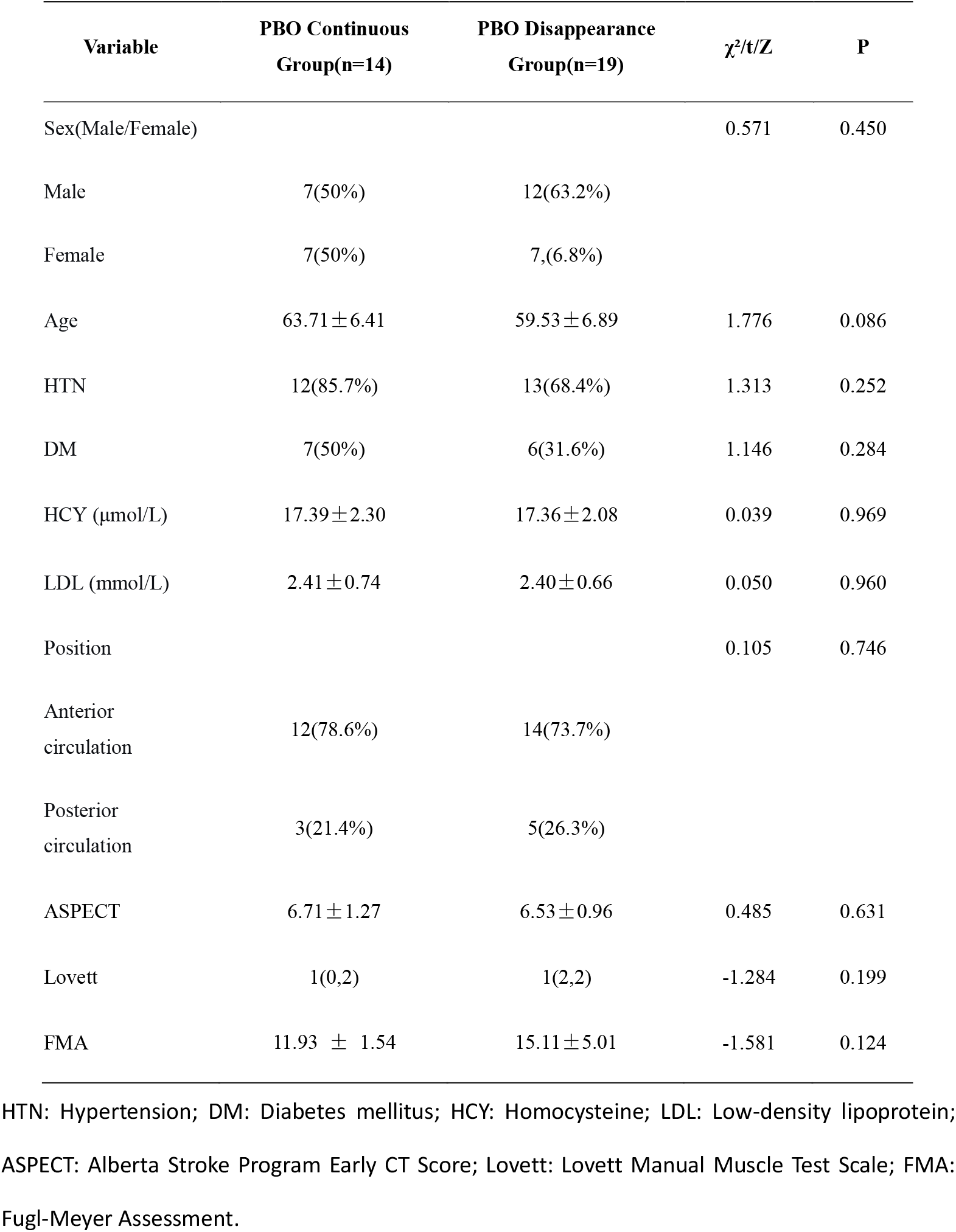
Comparison of Baseline Data Between the PBO Discontinuation Group and the PBO Continuation Group.

| Variable | PBO Continuous<br>Group(n=14) | PBO Disappearance<br>Group(n=19) | $\chi^2/t/Z$ | P |
| --- | --- | --- | --- | --- |
| Sex(Male/Female) |  |  | 0.571 | 0.450 |
| Male | 7(50%) | 12(63.2%) |  |  |
| Female | 7(50%) | 7,(6.8%) |  |  |
| Age | 63.71±6.41 | 59.53±6.89 | 1.776 | 0.086 |
| HTN | 12(85.7%) | 13(68.4%) | 1.313 | 0.252 |
| DM | 7(50%) | 6(31.6%) | 1.146 | 0.284 |
| HCY (μmol/L) | 17.39±2.30 | 17.36±2.08 | 0.039 | 0.969 |
| LDL (mmol/L) | 2.41±0.74 | 2.40±0.66 | 0.050 | 0.960 |
| Position |  |  | 0.105 | 0.746 |
| Anterior<br>circulation | 12(78.6%) | 14(73.7%) |  |  |
| Posterior<br>circulation | 3(21.4%) | 5(26.3%) |  |  |
| ASPECT | 6.71±1.27 | 6.53±0.96 | 0.485 | 0.631 |
| Lovett | 1(0,2) | 1(2,2) | -1.284 | 0.199 |
| FMA | 11.93 ± 1.54 | 15.11±5.01 | -1.581 | 0.124 |

HTN: Hypertension; DM: Diabetes mellitus; HCY: Homocysteine; LDL: Low-density lipoprotein; ASPECT: Alberta Stroke Program Early CT Score; Lovett: Lovett Manual Muscle Test Scale; FMA: Fugl-Meyer Assessment.

3. Comparison of Lovett Grading and FMA Between the PBO Disappearance Group and the PBO Persistence Group at 3 Months

After 3 months, the PBO-resolved group demonstrated significantly higher FMA scores than the non-PBO group (P < 0.05). No statistically significant difference was observed between the two Lovett grades (P > 0.05), as shown in Table 4.

**Table 4.** Comparison of Lovett Grading and FMA Scores at 3 Months Between the PBO Disappearance Group and the PBO Persistence Group.

|  | **PBO Continuous**<br>**Group(n=14)** | **PBO Disappearance**<br>**Group(n=19)** | **t/Z** | **P** |
| --- | --- | --- | --- | --- |
| LovettM(P25,P75) | 3(2,4) | 4(3,4) | -1.594 | 0.111 |
| FMA | 27.00±8.59 | 34.11±8.04 | -2.437 | 0.021 |

Lovett: Lovett Manual Muscle Test Scale; FMA: Fugl-Meyer Assessment.

## Discussion

Post-Brachial Oculomotor Phenomenon (PBO) represents a distinct and often overlooked clinical manifestation in stroke patients with hemiplegia, characterized by involuntary elevation of the paralyzed limb during yawning^[9]^. Among the 33 patients exhibiting PBO in this study, 31 demonstrated involuntary elevation solely in the upper limb, while 2 exhibited simultaneous elevation in both upper and lower limbs. Given this phenomenon, the study analyzed only upper limb function using the Lovett classification and FMA score. The Lovett classification is frequently used clinically for its intuitive assessment of limb functional changes. The FMA score quantifies upper limb motor ability through various movements, providing a more accurate evaluation of fine motor skills^[10]^. Yawning is a complex physiological behavior involving multi-level neural regulation: its initiation and control primarily rely on the paraventricular nucleus of the hypothalamus, which projects signals via oxytocinergic, cholinergic, and other neuronal groups to the hippocampus, brainstem (pons, medulla), and even the spinal cord^[11]^. Pathological or excessive yawning represents a clinically significant manifestation following acute stroke. Research indicates it frequently signals damage to the brainstem reticular formation or cortical/subcortical regions (particularly the insula and caudate nucleus). This demonstrates direct overlap between the neural pathways governing yawning and the lesion site in stroke^[12]^.

Due to the relatively low incidence of PBO, this study employed propensity score matching (PSM) to select patients with comparable baseline characteristics for comparison with the PBO cohort, ensuring research rigor^[13]^. A total of 33 PBO patients were recorded and matched 1:2 via PSM, resulting in 26 patients (PBO group) matched to 52 corresponding patients (non-PBO group). Further analysis revealed that at 3 months, the PBO group demonstrated statistically significant superiority over the non-PBO group in Lovett scores(Figure 1) and FMA scores (P < 0.05). The mechanism underlying PBO remains unclear, with three primary hypotheses proposed: 1. Cortical Inhibition Release Hypothesis: Following cortical injury (particularly in the internal capsule region, the most common lesion site for PBO), the inhibitory influence on subcortical motor structures (such as the brainstem reticular formation) diminishes. Yawning, as a potent physiological stimulus, may activate these “released” primitive pathways, triggering limb movements via structures like the reticulospinal tract^[3]^. 2. Affective-Motor System Hypothesis: Emotional states like drowsiness accompanying yawning may activate an “affective-motor system” independent of the pyramidal tract. This system stimulates oxytocinergic and cholinergic neuronal populations to release neurotransmitters such as dopamine, excitatory amino acids, and oxytocin. Its descending fibers directly excite brainstem and spinal motor neurons^[14]^; 3. Proprioceptive Loop Hypothesis: The intense contraction of respiratory muscles during yawning sends proprioceptive signals retrograde to the spinal cord anterior horn and cerebellum, ultimately reaching the lateral reticular nucleus of the medulla via the ventral cerebellar spinal tract. Subsequently, motor signals from the lateral reticular nucleus are generated in response and travel via the cerebellar extrapyramidal pathway back to anterior horn cells at C4 to C8, causing involuntary movements in paralyzed upper limbs(Figure 2). In this context, yawning is considered an externalization of a homeostatic arousal mechanism and is frequently observed post-stroke. A limitation of this theory is its inability to explain lower limb movements observed in certain case reports^[15,16]^. Regardless of the hypothesis, the occurrence of PBO strongly suggests that although patients have lost the corticospinal tract—the higher-level motor “highway”—their brainstem-spinal cord level primitive motor networks, such as the reticulospinal system and cerebellar-brainstem connections, remain relatively intact or in a state of heightened excitability. In this study, patients in the PBO group demonstrated significantly superior muscle strength and motor function scores at 3 months compared to the non-PBO group, suggesting that PBO serves as a clinical marker of the integrity and excitability of primitive motor pathways. The PBO phenomenon itself provides living proof that the lower-level motor centers in the brainstem and spinal cord retain functional integration capabilities and that neural connections to limb muscles remain partially intact. The ability to perform the complex coordinated movement of PBO indicates that the “alternative pathway” from the brainstem to the anterior horn cells of the spinal cord is structurally preserved and functionally activatable. Compared to non-PBO patients, those exhibiting PBO possess more complete “neurological reserve.” During post-stroke neural functional remodeling, these preserved, activatable primitive pathways provide a crucial anatomical and physiological foundation for motor recovery. Rehabilitation training may more readily compensate for impaired corticospinal tract function by strengthening and “taming” these proven pathways, thereby achieving superior functional outcomes.

**Figure 1:**
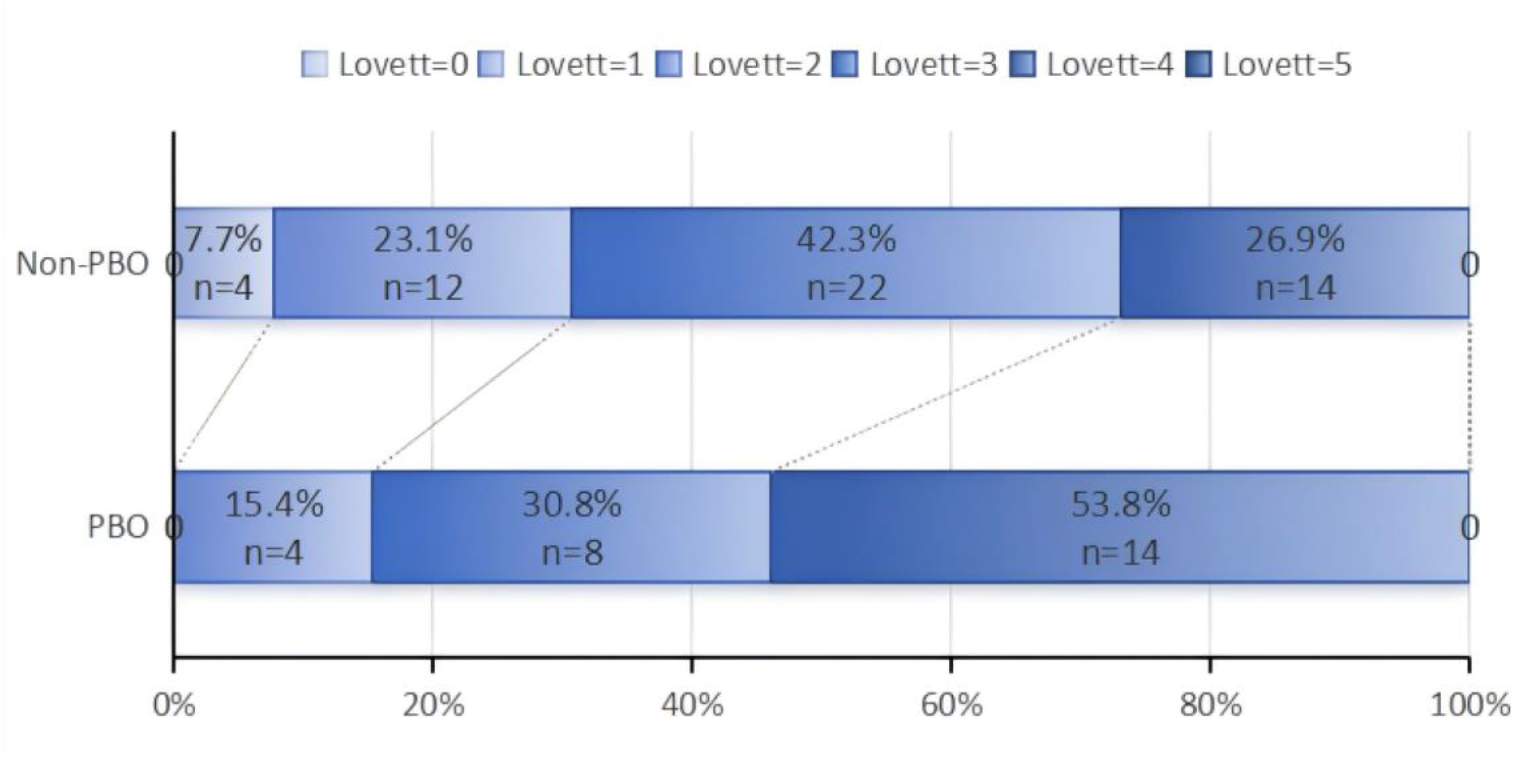
Lovett score at 3 mouths for all patients.

**Figure 2.**
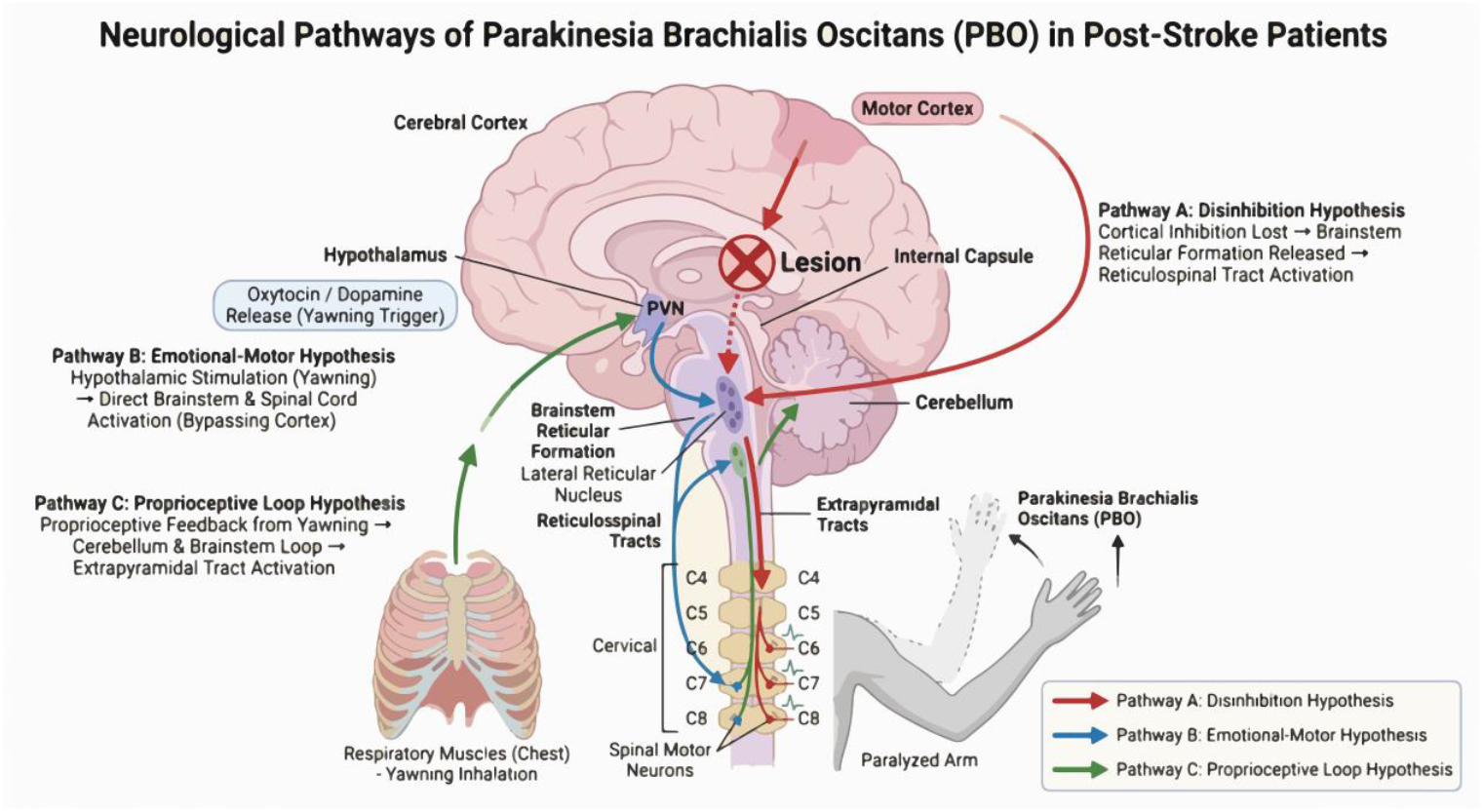

Literature reports suggest that PBO may be associated with poor patient outcomes^[9]^. We classified patients whose PBO resolved within one month post-stroke as the resolution group and those with persistent PBO as the persistence group. Statistical analysis of Lovett muscle strength grades and FMA scores between these groups revealed no statistically significant difference in Lovett muscle strength grades (P > 0.05). The FMA scores in the disappearance group were significantly higher than those in the persistence group (P < 0.05). In this study, the FMA score focuses more on evaluating isolated movements, coordination, and fine motor control, which are highly dependent on the integrity of the corticospinal tract^[17]^. The higher FMA scores in the disappearance group strongly suggest better recovery of corticospinal tract function. Regarding this outcome, we venture the hypothesis that the persistent PBO phenomenon may serve as a clinical marker of the motor system’s failure to transition successfully from a “lower-level compensatory mode” to a “higher-level dominant mode” post-stroke. This may indicate that neural remodeling has become stuck in an inefficient plateau phase. The superior function in the PBO-disappeared group likely stems not only from the “re-inhibition” of lower-level centers following CSM recovery but also from an active, successful “functional hierarchical transition” process. In the persistent PBO group, impaired recovery of the corticospinal tract leaves lower pathways persistently overactive and “unleashed” due to lack of higher-level control. This state itself may hinder corticospinal reorganization through mechanisms like synaptic competition, creating a vicious cycle^[18]^. Consequently, functional recovery remains stalled at a plateau, preventing further improvement.

This study reveals that the clinical significance of persistent biceps brachii movement (PBO) extends far beyond a simple “presence or absence” binary prognostic marker. The persistence of PBO should be regarded as a critical “clinical decision-making inflection point.” For these patients, rehabilitation strategies must shift from universal training to intensive, targeted interventions focused on “disinhibition and recontrol.” The early disappearance of PBO strongly indicates successful remodeling of corticospinal tract function and the “return” of higher-level motor control, signaling that the nervous system is progressing toward a more optimal functional reorganization. For these patients, the timing and quality of rehabilitation intervention are critical. Around the time window of PBO resolution, delivering high-intensity, precision rehabilitation focused on promoting cortical control and optimizing movement patterns may yield disproportionately high returns, thereby maximizing their functional recovery potential.

This study has several limitations. First, it is observational in nature. Although propensity score matching was employed to reduce confounding bias, the sample size—particularly in the PBO group—remains relatively small. This may have impacted statistical power and precluded in-depth subgroup analyses based on specific lesion locations (e.g., internal capsule, pons, subcortical regions), thereby limiting the generalizability of conclusions and the exploration of mechanisms across different injury patterns. Second, functional outcomes were primarily assessed using clinical scales, lacking direct neuroimaging and electrophysiological evidence. The intrinsic mechanisms linking PBO to functional recovery at the neural circuit level remain unexplored. Future research aims to employ multimodal neuroinformation fusion (synchronous acquisition and analysis of high-resolution MRI, DTI, high-density EEG/motor-related potentials, and surface EMG) to quantitatively analyze PBO-specific neural circuit characteristics in an expanded sample. Longitudinal observation of their dynamic evolution will provide more robust evidence for understanding post-stroke motor function remodeling.

## Data availability statement

The raw data supporting the conclusions of this article will be made available by the authors without undue reservation. The original contributions presented in this study are included in the article/supplementary material. Further inquiries can be directed to the corresponding author(s).

## Ethics statement

This study was conducted in accordance with the Declaration of Helsinki (as revised in 2013).

The study protocol was reviewed and approved by the ethics committee of The First Affiliated Hospital of Henan Medical University (IRB approval number: EC-024-588) and conformed to the ethical standards for medical research involving human subjects. Participants provided written informed consent prior to taking part in the study.

## Author contributions

Congcong Wang, Runying Wang and Hua Hu were responsible for the conception and design of the article, collection and organization of data, statistical processing, composition, and revision of the thesis. Shuangxi Guo carried out the implementation and feasibility analysis of the research, was in charge of the quality control and proofreading of the article, and was overall responsible for, supervising, and managing the article.Xiaojun Tian and Zhou Su were involved in the conception and design of the article and the revision of the thesis.

## References

1. Ma Q, Li R, Wang L, et al. Temporal trend and attributable risk factors of stroke burden in China, 1990-2019: an analysis for the global burden of disease study 2019. Lancet Public Health. 2021;6(12):e897–e906. doi:10.1016/S2468-2667(21)00228-0

2. Nave AH, Rackoll T, Grittner U, et al. Physical fitness training in patients with subacute stroke (PHYS-STROKE): multicentre, randomised controlled, endpoint blinded trial. BMJ. 2019;366:5101. doi:10.1136/bmj.l5101

3. Walusinski O, Quoirin E, Neau JP. [parakinesia brachialis oscitans]. Rev Neurol (Paris). 2005;161(2):193–200. doi:10.1016/s0035-3787(05)85022-2

4. Lim ICZY, Neo S. Parakinesia brachialis oscitans: old sign, new findings. Stroke. 2022;53(2):e60–e62. doi:10.1161/STROKEAHA.121.037124

5. Kleindorfer DO, Towfighi A, Chaturvedi S, et al. 2021 guideline for the prevention of stroke in patients with stroke and transient ischemic attack: a guideline from the american heart association/american stroke association. Stroke. 2021;52(7):e364–e467. doi:10.1161/STR.0000000000000375

6. Naganuma M, Tachibana A, Fuchigami T, et al. Alberta stroke program early CT score calculation using the deep learning-based brain hemisphere comparison algorithm. J Stroke Cerebrovasc Dis Off J Natl Stroke Assoc. 2021;30(7):105791. doi:10.1016/j.jstrokecerebrovasdis.2021.105791

7. Kniep HC, Elsayed S, Nawabi J, et al. Imaging-based outcome prediction in posterior circulation stroke. J Neurol. 2022;269(7):3800–3809. doi:10.1007/s00415-022-11010-4

8. Wang Z, Zhang T, Fan J, et al. Clinical validation of automated depth camera-based measurement of the fugl-meyer assessment for upper extremity. Clin Rehabil. 2024;38(8):1091–1100. doi:10.1177/02692155241251434

9. Chowdhury A, Datta AK, Biswas S, Biswas A. Parakinesia brachialis oscitans - a rare post-stroke phenomenon. Tremor Hyperkinetic Mov N Y N. 2022;12:6. doi:10.5334/tohm.680

10. Wiesner K, Schwarz A, Meya L, et al. Interrater reliability of the fugl-meyer motor assessment in stroke patients: a quality management project within the ESTREL study. Front Neurol. 2024;15:1335375. doi:10.3389/fneur.2024.1335375

11. Wani PD, Agarwal M. The science of yawning: exploring its physiology, evolutionary role, and behavioral impact. J Fam Med Prim Care. 2025;14(8):3115–3120. doi:10.4103/jfmpc.jfmpc_1677_24

12. Teive HAG, Munhoz RP, Camargo CHF, Walusinski O. Yawning in neurology: a review. Arq Neuropsiquiatr. 2018;76(7):473–480. doi:10.1590/0004-282X20180057

13. Austin PC. Optimal caliper widths for propensity-score matching when estimating differences in means and differences in proportions in observational studies. Pharm Stat. 2011;10(2):150–161. doi:10.1002/pst.433

14. de Lima PMG, Munhoz RP, Becker N, Teive HAG. Parakinesia brachialis oscitans: report of three cases. Parkinsonism Relat Disord. 2012;18(2):204–206. doi:10.1016/j.parkreldis.2011.09.020

15. Moritz C, Field-Fote EC, Tefertiller C, et al. Non-invasive spinal cord electrical stimulation for arm and hand function in chronic tetraplegia: a safety and efficacy trial. Nat Med. 2024;30(5):1276–1283. doi:10.1038/s41591-024-02940-9

16. Walusinski O, Neau JP, Bogousslavsky J. Hand up! Yawn and raise your arm. Int J Stroke Off J Int Stroke Soc. 2010;5(1):21–27. doi:10.1111/j.1747-4949.2009.00394.x

17. Rowe V, Blanton S, Aycock D, Hayat MJ, Ali SZ. Remote delivery of the fugl-meyer assessment for the upper extremity: a pilot study to assess feasibility, reliability, and validity. Arch Rehabil Res Clin Transl. 2023;5(2):100261. doi:10.1016/j.arrct.2023.100261

18. Mason J, Frazer A, Horvath DM, et al. Adaptations in corticospinal excitability and inhibition are not spatially confined to the agonist muscle following strength training. Eur J Appl Physiol. 2017;117(7):1359–1371. doi:10.1007/s00421-017-3624-y

